# Prescribed Cardiac Wearables in Routine Care: a qualitative study of Patient Experiences

**DOI:** 10.64898/2026.04.09.26350550

**Authors:** Aileen Zeng, Edel O’Hagan, Ritu Trivedi, Belinda Ford, Timothy Perry, Samual Turnbull, Brodie Sheahen, John Mulley, Maha Sedhom, Carina Choy, Adrian Biasi, Scott Walters, J. Jaime Miranda, Clara K Chow, Liliana Laranjo

## Abstract

**Background:** Continuous adhesive patch electrocardiographic (ECG) wearables are increasingly prescribed. Patient experience with these devices can influence adherence, but research in this area is limited. This study aimed to explore the perceptions and experiences of patients receiving wearable cardiac monitoring technology as part of their routine care through the lens of treatment burden.

**Methods:** This was a qualitative study with semi-structured phone interviews conducted between February and May 2024. We recruited participants from primary care and outpatient clinics using maximum variation sampling to ensure diversity in sex, ethnicity, and education levels. Interviews were audio-recorded, transcribed, and analysed using reflexive thematic analysis.

**Results:** Sixteen participants (mean age 51 years, 63% female) were interviewed (average duration: 33 minutes). Three themes were developed: 1) ‘Experience using the device: Burden vs Ease of Use’, which captured participants’ perceptions of how easily they could integrate the device in their daily lives; 2) ‘Individual variability in responses to ECG self-monitoring’ covered participants’ emotional and cognitive response to knowing their heart rhythm was monitored; and 3) ‘The care process shapes patient experiences’ reflected support preferences during the set-up and monitoring period and the uncertainty regarding timely clinical and device feedback.

**Conclusions:** Patients valued cardiac wearables for facilitating diagnosis and felt reassured knowing they were clinically monitored. However, gaps in information provided to patients seemed to cause anxiety for some participants. These concerns could be mitigated through clearer clinician communication and patient education at the time of prescription.

## Introduction

Cardiac investigations, including an electrocardiogram (ECG), play a key role in diagnosing whether symptoms (e.g., palpitations, syncope, dyspnoea, chest pain) are benign or a sign of underlying cardiovascular condition(s).^1^ Traditional ECG monitoring methods, such as the Holter monitor, are typically deployed for 24 hours and have limited utility for detecting intermittent or sporadic symptomatic events.^2^ Longer-term continuous ECG monitoring can detect heart rhythm abnormalities that would otherwise be missed by traditional 24-hr monitoring.^1,3–5^

Long-term continuous ambulatory ECG monitors, such as patch monitors, can be connected to smartphone apps to aid user engagement and incorporate near-real-time analytics by transmitting data to a central monitoring platform.^6^ Since 2020, patch ECG monitors have gained wider acceptance, with clinicians increasingly adopting point-of-care investigations outside traditional settings to enable faster diagnoses.^5,7,8^ Prescription of a patch ECG monitor depends on availability, frequency of symptoms (weekly or monthly), patient preference and digital health literacy, as part of the shared decision-making process.^1^

Understanding patient experiences with prescribed wearables informs what is critical to successful patient engagement and future adherence. Existing studies primarily focus on consumer devices (i.e., Fitbits).^9–11^ Given the novelty of the technology and its application in clinical care, qualitative inquiry is needed to understand engagement with medical-grade devices, particularly in relation to how digital competency interacts with usability. These factors can limit engagement with monitoring and hamper arrhythmia detection in real-world settings.^1,12,13^ The patient’s perception of the cumulative workload of healthcare tasks, including investigations, is often referred to as treatment burden,^14^ and may reduce engagement with medical advice.^15^ Insights from patients’ lived experiences enable healthcare providers to identify the support needed and highlight gaps that hinder the integration of digital devices into patient care.^16^

This qualitative study aimed to explore the perceptions and experiences of patients receiving wearable cardiac monitoring technology as part of their routine care through the lens of treatment burden.

## Materials / Methods

This qualitative study, with semi-structured interviews, explored patients’ lived experience with using a prescribed patch ECG monitor. Phenomenology, defined as a theoretical approach to exploring the meaning of individuals’ perspectives on a phenomenon, underpinned the methodological orientation.^17,18^ This study used the Consolidated Criteria for Reporting Qualitative Research (COREQ) research checklist.^19^ for guidance when reporting this manuscript (Supplement 1).

The human research ethics committee at Western Sydney Local Health District granted ethics approval (2022/ETH00269). All participants provided informed consent before participating in the qualitative interviews, with consent forms signed through REDCap (Research Electronic Data Capture). Data were stored on secure servers, and access was restricted to approved study personnel. We de-identified data (e.g. age and name) before transcription and analysis.

The analysis was conducted by AZ, a podiatrist with no prior relationship to study participants. Other team members included a non-practising physiotherapist (EO), a general practitioner (LL), a cardiologist (CC) and a research fellow (RT), all with experience in digital health and cardiovascular care. The primary author has a research interest in treatment burden. These professional backgrounds may have influenced interpretation, and reflexivity was maintained throughout the analysis.

Eligible participants, adults receiving a patch ECG monitor (HeartBug™) as part of their routine care, were recruited from twenty-four General Practice (GP) clinics in Western Sydney and an outpatient arrhythmia clinic at a tertiary hospital in Sydney. Leaflet advertisements with a QR code linking to a patient consent form were distributed by clinicians (including nurses, doctors, and pharmacists) when patients attended their first follow-up post-discharge. Patients were handed a leaflet inside the Heartbug box to provide information about the study and facilitate recruitment.^20^ (Detailed description of the settings in Supplement 2). Participants were recruited during the implementation of a broader government-funded healthcare improvement initiative that provided eligible patients with free access to HeartBug™ devices. Eligible patients, from both settings, were investigated for episodes of intermittent cardiac arrhythmias or episodes of syncope or had suspected or were at risk of a cerebrovascular accident. We used maximum variation sampling to identify and recruit participants representing diverse sex, ethnicities, and education levels. Participants had an option to involve a translator if English was not their preferred language. Participants received a $25 voucher upon completing the interview.

The HeartBug™ is a prescribed, smartphone-connected single-lead adhesive patch electrocardiographic monitor that can be worn for up to 28 days.^21^ The device is small (i.e., 3cm in diameter), non-invasive, and waterproof. Data are transmitted in two directions: 1) from the device to the smartphone via Bluetooth, and 2) from the smartphone to the central monitoring unit via the internet. Transmitted ECG segments include descriptive annotations (e.g. time) from the app. Participants can log their symptoms (e.g., palpitations) by pressing a “record button” in the smartphone app, enabling clinicians to correlate reported symptoms with recorded ECG data. Alternatively, the device records the ECG in short segments when it detects an abnormal rhythm.

A cardiologist asynchronously analyses the recording at the end of the monitoring period and sends a report to the prescribing clinician, as part of an Australian Government-reimbursed service.^22^ Participants were instructed to always wear the device (including showering) for up to 28 days (duration varying depending on clinical advice).^23^

The interview guide (Supplement 3) was pilot tested within the research team. One researcher (AZ) conducted 1:1 semi-structured telephone interviews, and audio files were transcribed verbatim using a transcription service and verified for accuracy. Interviews were conducted until the researchers deemed sufficient data had been collected to generate themes that answered the research question.^17,24^

We used a six-phase reflexive thematic analysis to analyse interview data (details on reflexive analysis in Supplement 4).^25^ We conducted inductive coding, transforming the data into statements representing meaning units (words/phrases that convey a single meaning), and then grouped codes based on thematic similarities in NVivo.^26^. The theoretical flexibility of thematic analysis, informed by phenomenology, enabled us to delve into patients’ perspectives on their experiences with the device and potential factors contributing to treatment burden. During the analysis, we applied the lens of treatment burden, guided by concepts from Al Zahidy’s taxonomy of digital treatment burden.^15^

## Results

Of the 42 expressions of interest, 16 participants were invited and interviewed (with an average duration of 33 minutes) between February and May 2024. The remaining 26 were not interviewed due to unsuccessful contact attempts or as a result of maximum variation sampling to ensure diversity. Nine participants were recruited from primary care (i.e. GP) and 7 from outpatient clinics. The mean age was 51±15 years; 10 (62.5%) were female, 11 (68.8%) were White, and 6 (37.4%) had secondary education or lower (Table 1). Three main themes and subthemes are described below. Tables 2 and 3, Figure 1 and Supplement 5 summarise themes, subthemes, and illustrative quotes. Supplement 6 summarises key considerations for implementing patch ECG monitors in clinical practice and outlines recommendations for educational materials informed by our qualitative findings.

**Figure 1.**
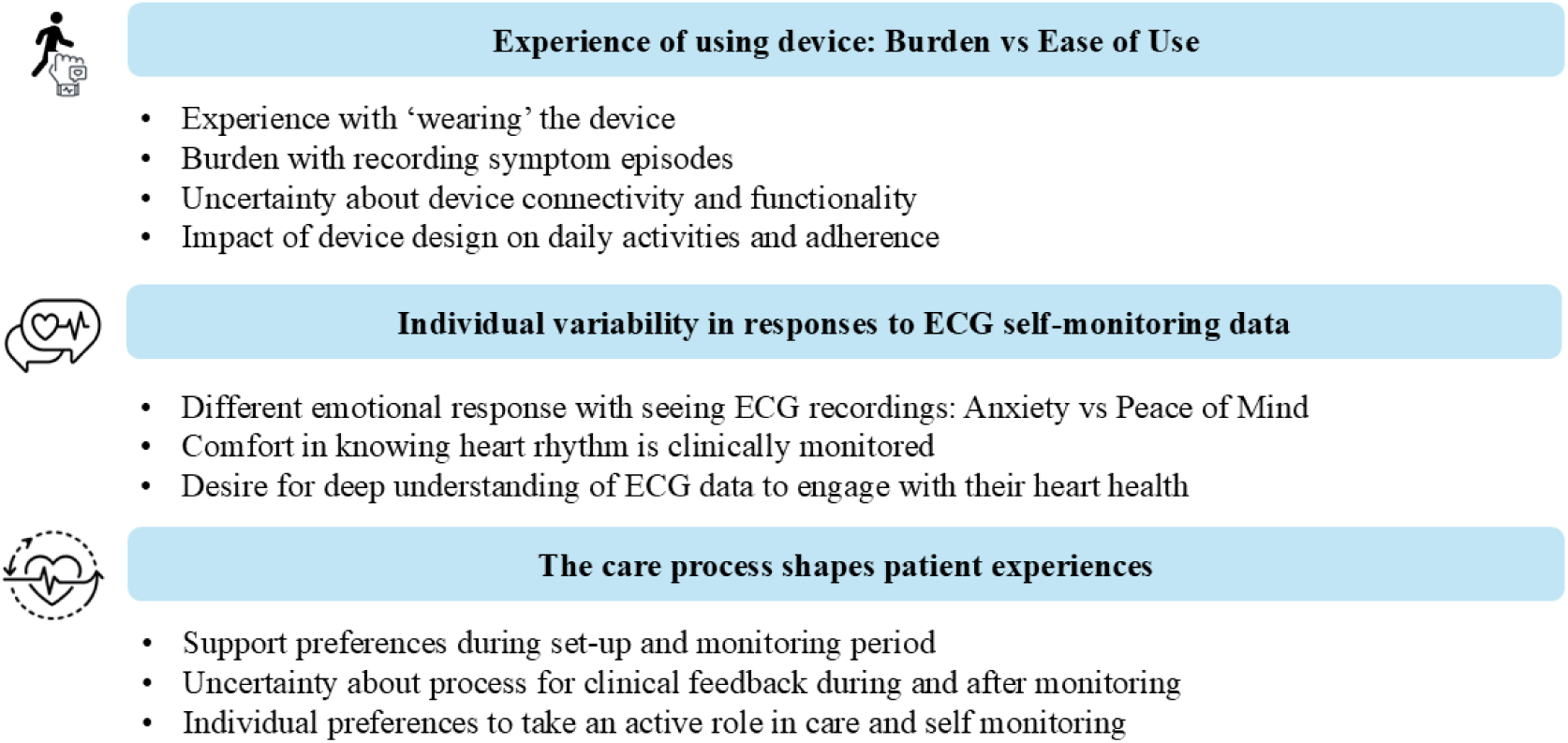
Themes and subthemes.

**Table 1.**
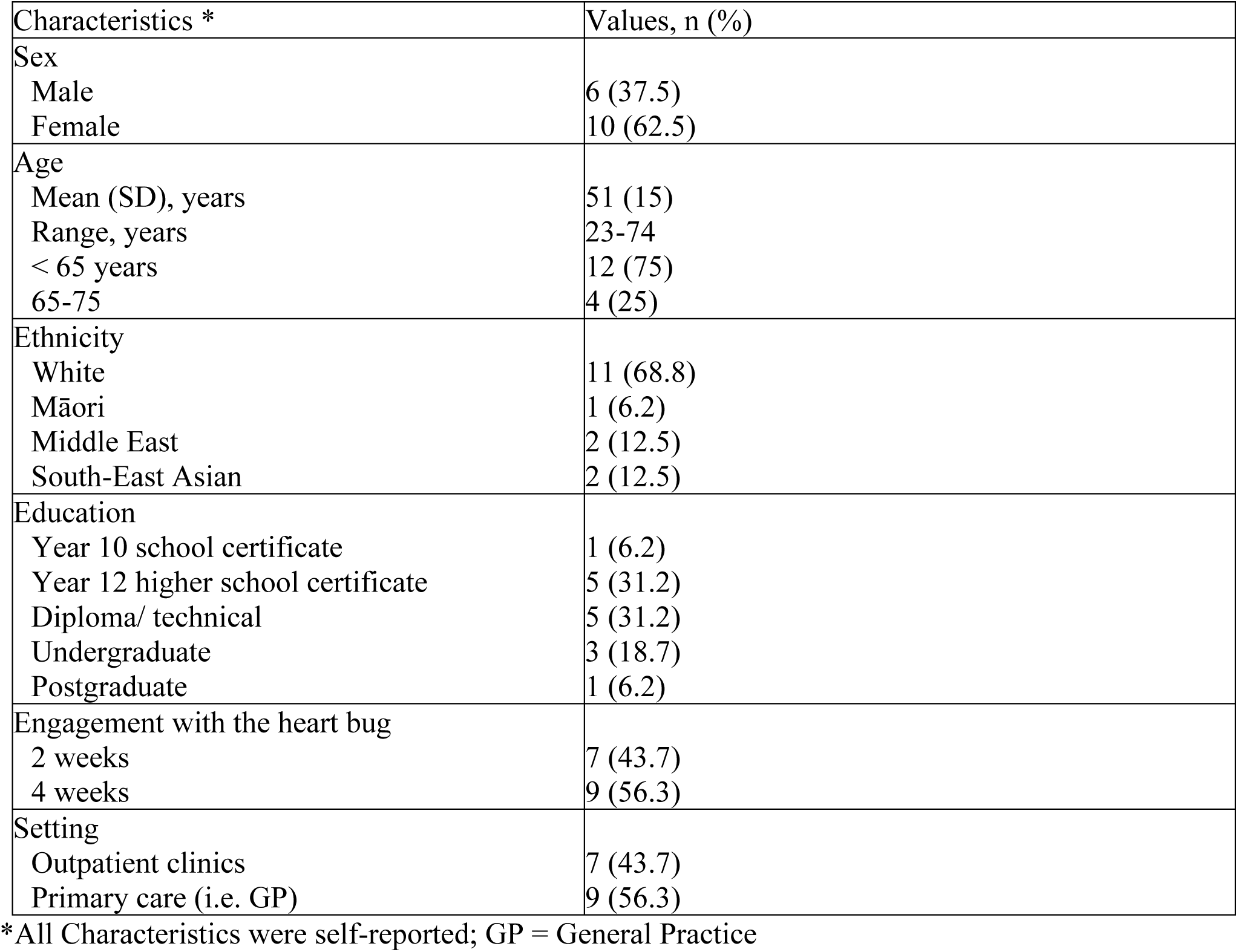
Participant characteristics and Engagement metrics.

**Table 2:** Main Themes From Interviews.

| Theme | Description |
| --- | --- |
| Experience of using the device: Burden vs Ease of Use | User lived experience with wearing the device as part of their daily life |
| Individual variability in responses to ECG self-monitoring data | Individuals' interactions with their ECG data on the app |
| The care process shapes patient experiences | How service delivery from clinical providers and tech company play a key role to the patient's overall experience with being prescribed and wearing with the device |

**Table 3:**
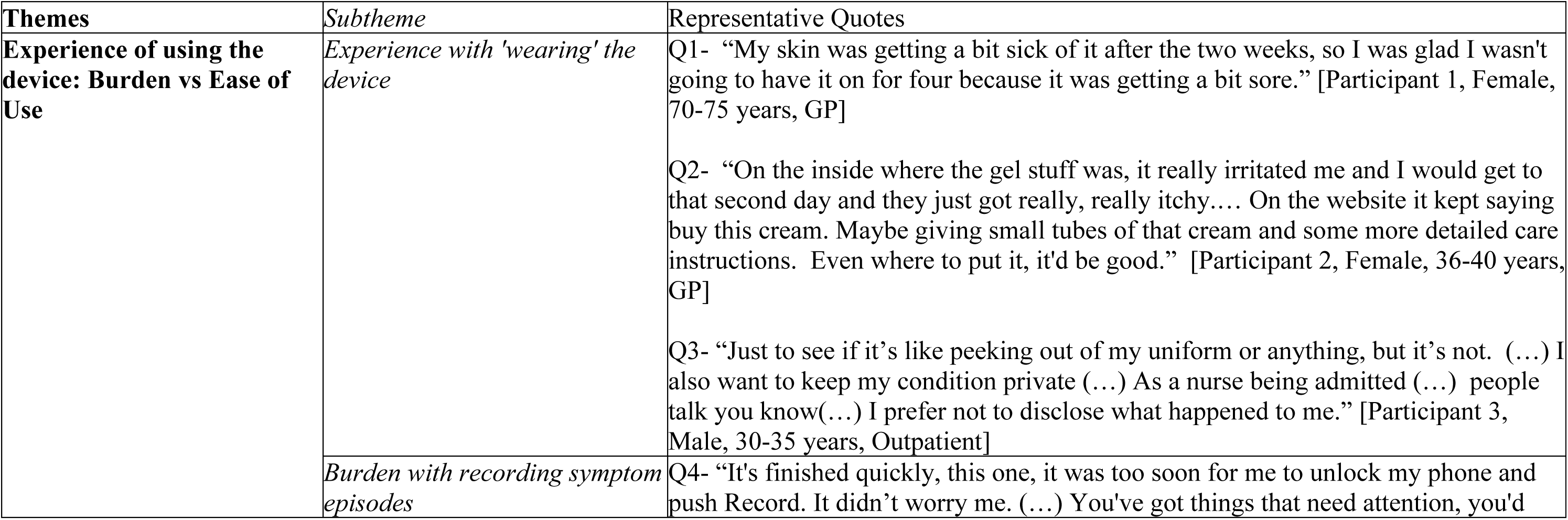
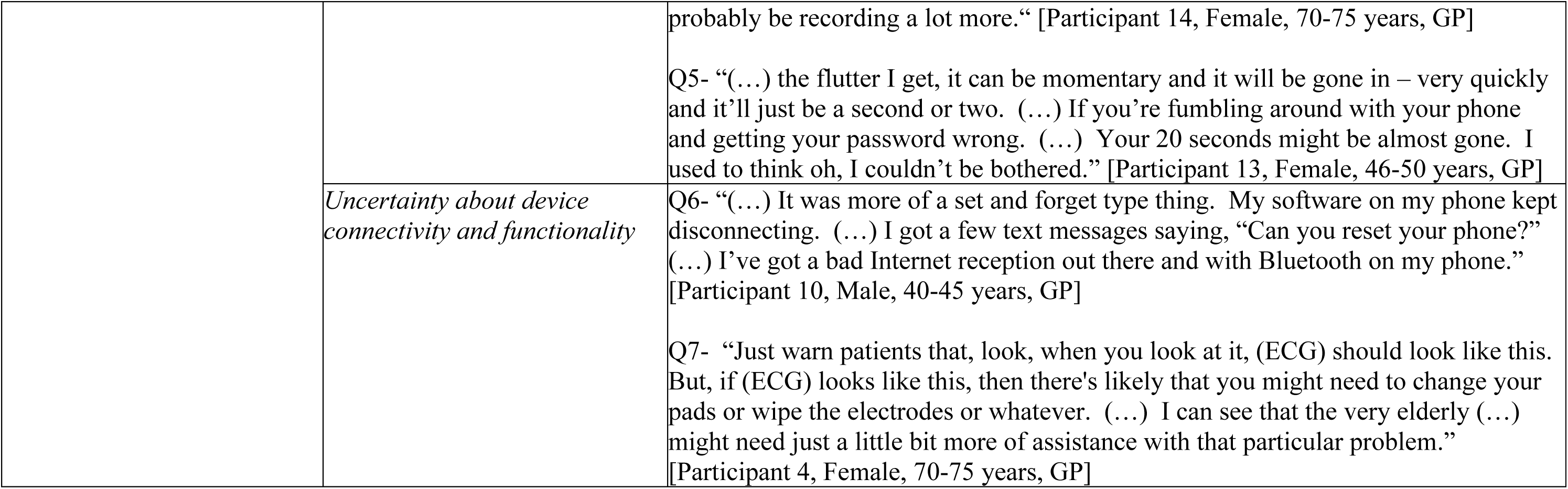
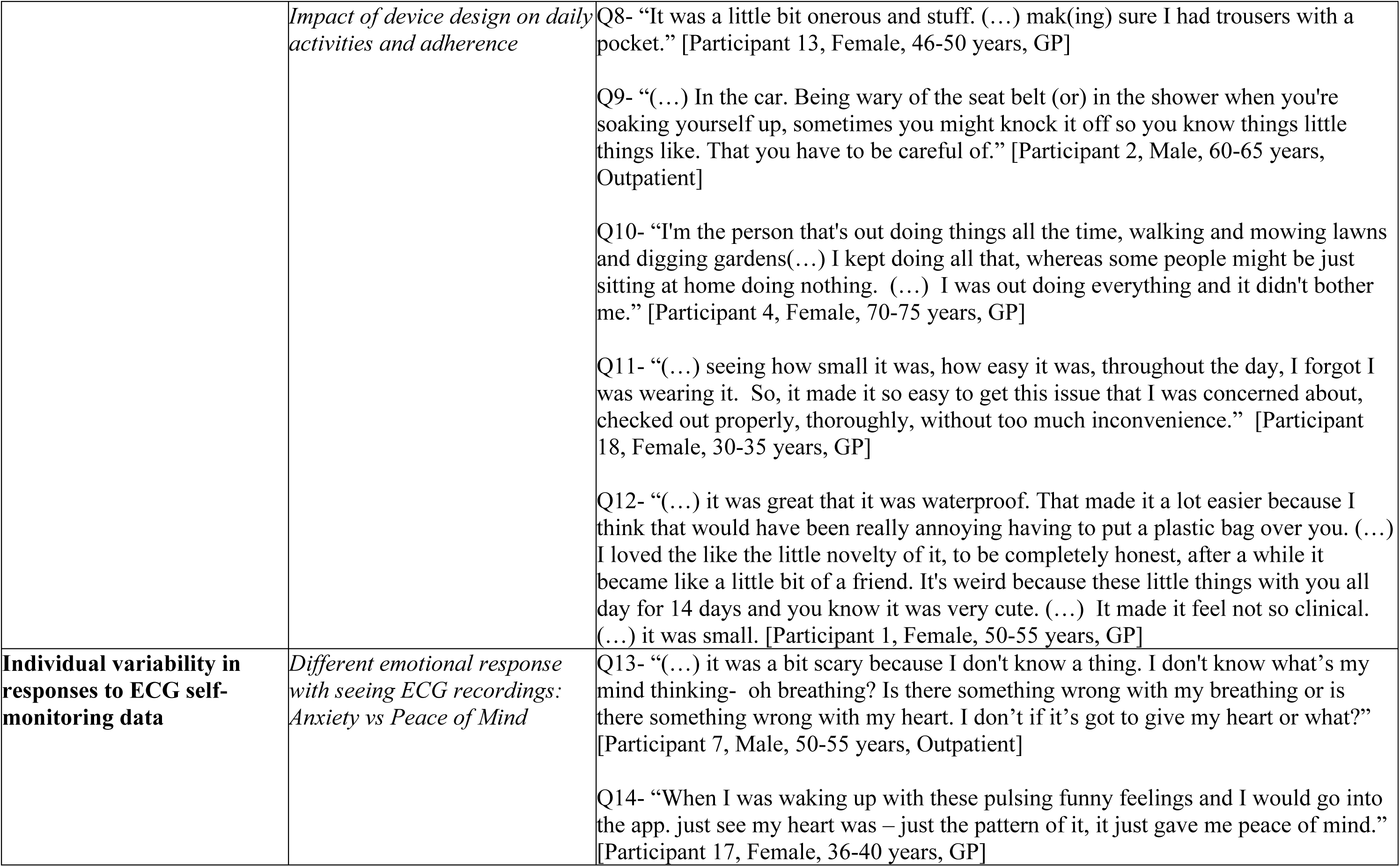
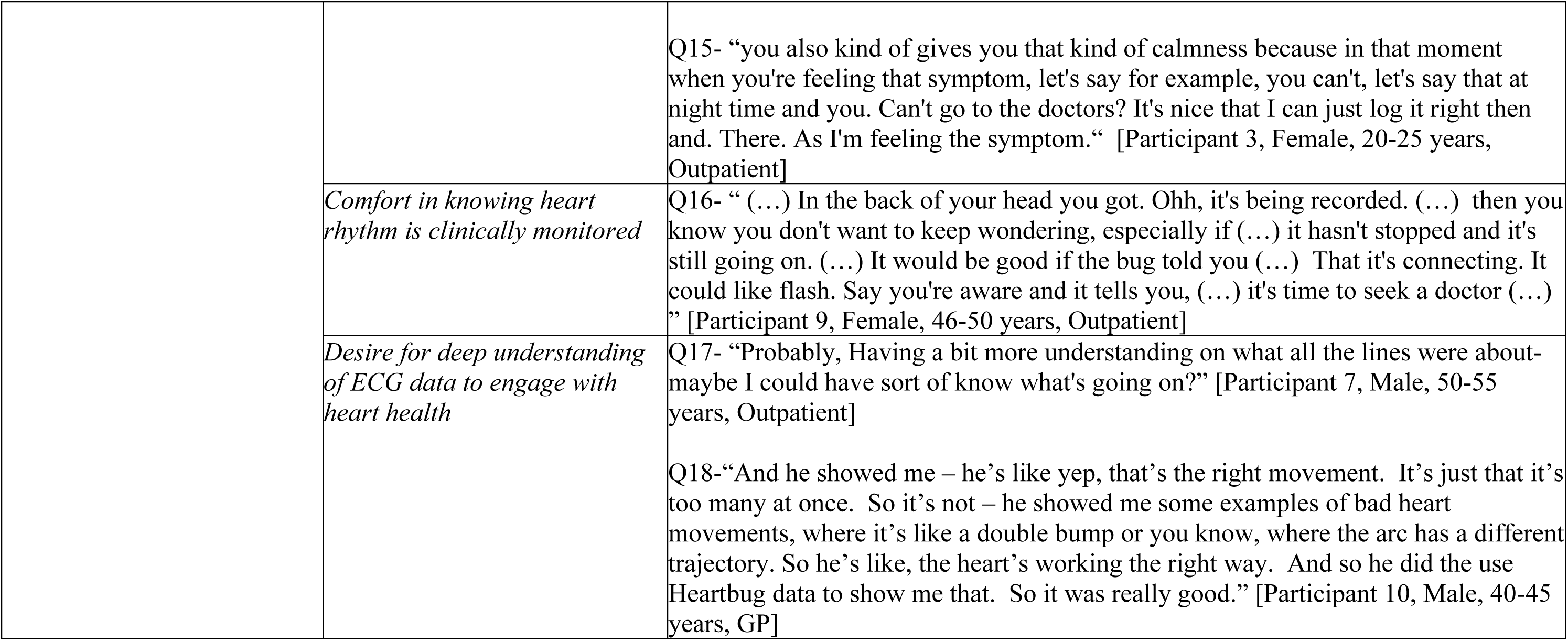
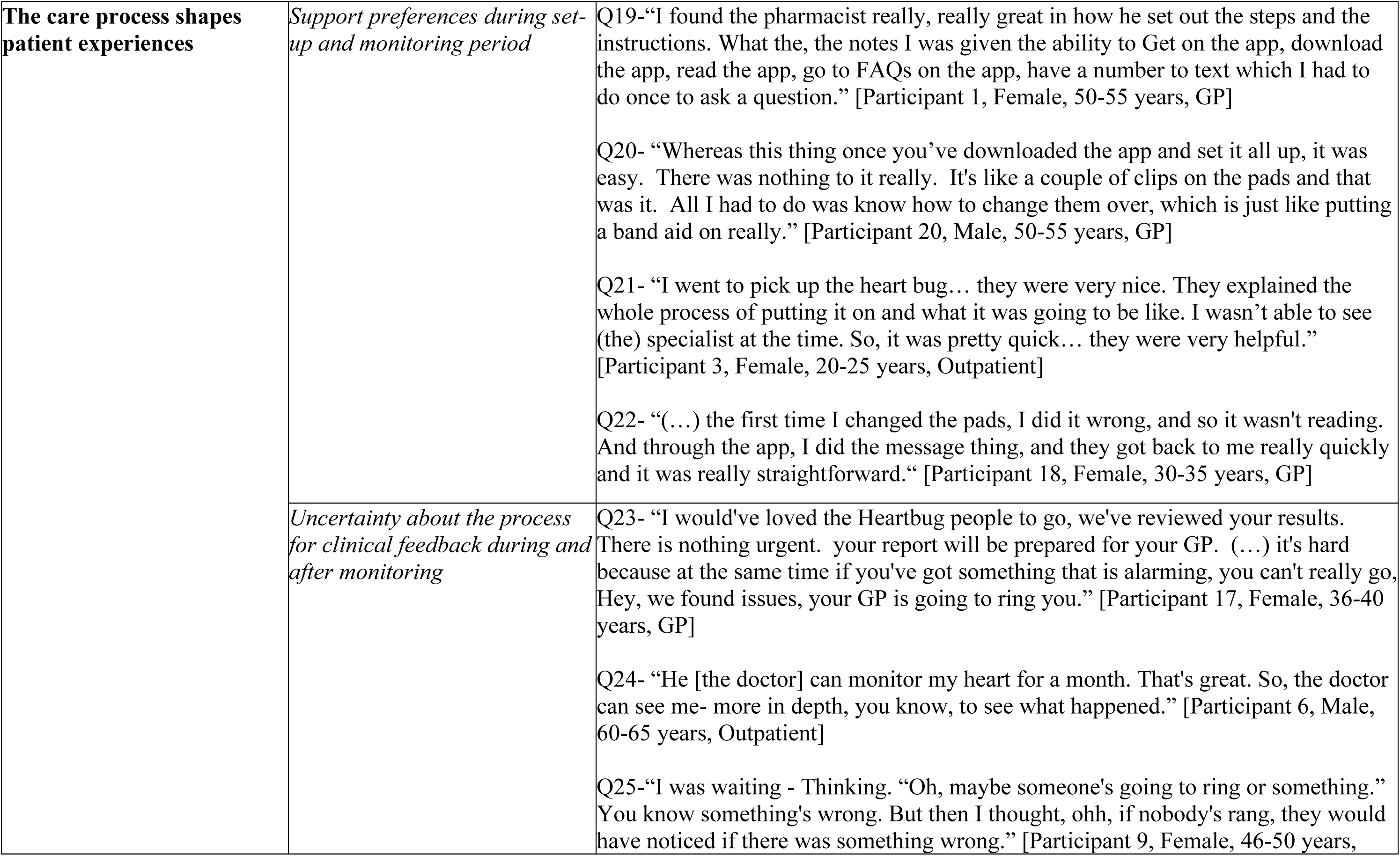
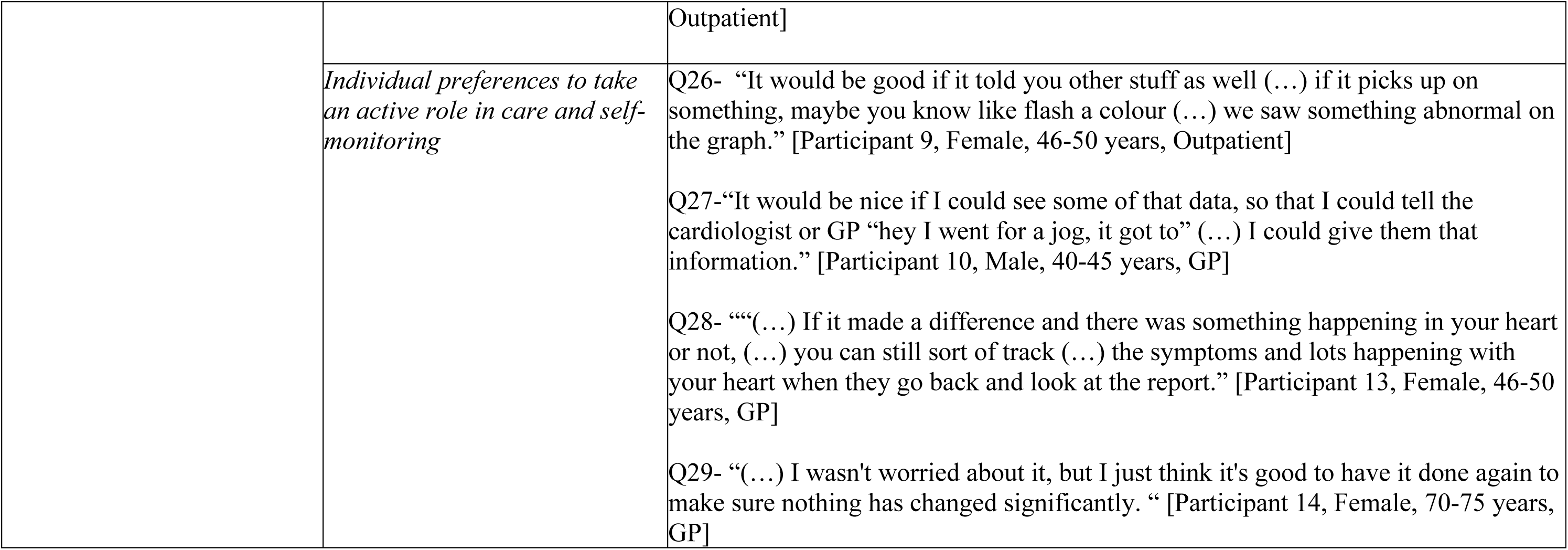
Illustrative Quotes for Themes and Subthemes.

### Experience of using the device: Burden vs Ease of Use

#### Experience with ’wearing’ the device

Participants expressed frustration with skin irritations, including itchiness and scarring, from the adhesive used in the electrode stickers, which contributed to an increased burden (Table 3, Quote 1). To mitigate these barriers to adherence, some participants learned to adjust the placement of the stickers or replace them. Some participants expressed the expectation that they would have been provided more details at the beginning about a spray that could have alleviated these skin irritations (Table 3, Quote 2).

Participants who wore the device for a longer duration (i.e., 4 weeks) reported social discomfort when it was visible. This visibility affected their social life because the device drew attention from others, prompting queries about the device or the user’s health. Some participants reported choosing clothing that could conceal the device for those reasons (Table 3, Quote 3).

#### Burden with recording symptom episodes

Some participants understood it was their responsibility to document symptoms yet also acknowledged the practical limitations and administrative burden associated with it (Table 3, Quote 4). Although many demonstrated the initiative to record symptoms via the app, some hesitated, either assuming the episode would be automatically recorded or feeling unmotivated to log the event during brief episodes (Table 3, Quote 5).

#### Uncertainty about device connectivity and functionality

Participants expressed uncertainty regarding whether their devices were successfully connected to their app, especially when a blank ECG screen appeared or the device emitted blinking signals (Table 3, Quote 6). Some participants expressed frustration that more information about potential troubleshooting issues was not provided earlier (Table 3, Quote 7). This was seen as a contextual factor that exacerbated the burden of maintaining the device. Those who wore the device for 4 weeks reported synchronisation issues related to the device’s Bluetooth connectivity and the position of the leads.

#### Impact of device design on daily activities and adherence

Participants had mixed responses about interferences during daily activities. Since the device required a Bluetooth connection to the app, participants forgot the necessity of keeping their phone in proximity. Some participants reported adapting by wearing clothing with pockets to carry their phones throughout the day (Table 3, Quote 8). Additionally, some participants reported that the electrodes could easily dislodge from the device, which would interrupt sleep or driving (because their position aligns with a seatbelt strap) (Table 3, Quote 9). Other participants reported that the device was easy to integrate into their daily activities. They noted that the device did not disrupt their work commitments or parental duties and that they continued with their usual active lifestyle, such as gardening (Table 3, Quote 10). Participants noted that the device’s benefits included its small size and waterproof feature. These features enabled patients to continue with daily activities (i.e. showering) and conceal them under clothing (Table 3, Quote 11). Some participants shared that the design made the device feel less clinical and associated it with being like a “little friend” (Table 3, Quote 12).

### Individual variability in responses to ECG self-monitoring data

#### Different emotional responses to seeing ECG recordings: Anxiety vs Peace of Mind

Some participants reported anxiety when viewing the ECG recordings on their app. Participants acknowledged limitations to their understanding of the diagnostic recordings, noting this could impact their emotional response (Table 3, Quote 13). However, other participants found that viewing the movement of the live ECG waveform provided reassurance that the device was functioning correctly (Table 3, Quote 14). Some participants noted that access to individual real-time ECG recordings provided emotional reassurance during periods of symptoms, stress, or uncertainty. Direct access to ECGs via the app offered acknowledgement or insight into symptoms they have experienced, and this was deemed valuable during times when clinical guidance was not immediately available (Table 3, Quote 15).

#### Comfort in knowing the heart rhythm is clinically monitored

Some participants felt ’peace of mind’ and alleviation of persistent concerns about their heart while wearing the device, and this reassurance was reinforced by knowing that their symptom episodes would be continuously monitored. They noted that having an ECG recording provided a pathway to diagnosis, which, in turn, reassured them that they would receive appropriate clinical attention (Table 3, Quote 16).

#### Desire for a deeper understanding of ECG data to engage with their heart health

Some participants felt curious when viewing their ECG recordings, particularly the waveform patterns, and were interested in understanding how these patterns visually changed under different scenarios (e.g., stressful periods) on the app (Table 3, Quote 17). Other participants engaged in discussions with their clinicians about ECG recordings, seeking to learn the difference between an abnormal ECG and a normal sinus rhythm (Table 3, Quote 18).

### The care process shapes patient experiences

#### Support preferences during the set-up and monitoring period

Some participants believed that having a facilitator for the initial device setup or receiving clearer information could boost their confidence in using the device and make it easier to learn (Table 3, Quote 19). However, not all participants believed in-person support was necessary, and some were confident they could navigate the application independently (Table 3, Quote 20). Information on the app, device functionality and features, a Q&A section, and advice on appropriate actions to take in the event of symptoms were most important to participants.

Participants found it helpful when education at setup covered the entire process of applying the device and what to expect. They also valued the quick setup and the fact that they did not need to rely on a specialist for assistance. This eased the technical and logistical aspects of device management. (Table 3, Quote 21). Participants found technical support useful for device maintenance (e.g. replacing electrode stickers or adjusting electrode placement to improve ECG signal quality) (Table 3, Quote 22).

#### Uncertainty about the process for clinical feedback during and after monitoring

Participants expressed uncertainty about how long they should wait for follow-up on their results. Most participants reported a preference for feedback within a couple of business days of completing their GP’s investigation, but acknowledged that clinical urgency could affect the timing (Table 3, Quote 23). Some outpatient participants reported an awareness that their results would be discussed at a scheduled follow-up appointment with their cardiologist, but were unclear whether results would be communicated to their GP (Table 3, Quote 24). After experiencing a symptom, some participants reported feeling scared or anxious awaiting clinician contact or follow-up. Participants trusted that their clinician would notify them if any clinically urgent episodes were detected, even if they did not log their symptoms in the app (Table 3, Quote 25).

#### Individual preferences to take an active role in care and self-monitoring

Participants reported individual preferences regarding the type and extent of device feedback they would like to receive. Some participants expressed interest in the app or device that alerts them to ECG abnormalities (Table 3, Quote 26). Some additional features they suggested included a notification to alert them when an anomaly is detected, or the ability to report on daily activities and add diary entries to support patient-clinician conversations (Table 3, Quote 27). Participants reported they understood the device was an integral component of the diagnostic process for their cardiac concerns, particularly the value of data collection through making personalised notes of their symptoms or anomalies in an app (Table 3, Quote 28). There were differing perceptions on the value of their role in tracking symptoms. Some participants expressed queries regarding the extent to which their collected data would influence their clinical management (Table 3, Quote 29). The device was often viewed as a mechanism to validate a health concern, such as facilitating a diagnosis, rather than as a collaborative tool for ongoing health engagement. Participants understood that extended monitoring periods facilitated greater data collection than conventional ECG monitoring.

## Discussion

### Main Findings

This study explored how patients experienced using a patch ECG monitor for extended cardiac monitoring to support clinical investigations. A common topic across themes and subthemes was the need for patient education on what to expect with the device. Overall, patients valued the use of these devices to facilitate clinical monitoring and saw them as an appropriate pathway to diagnosis, despite reporting barriers with skin irritation and logging symptom episodes. While participants demonstrated an interest in greater involvement in their care, there were mixed views about how direct access to ECG data could provide emotional reassurance or exacerbate patient anxiety. Interpreted through the lens of treatment burden, these findings suggest that uncertainty and additional health-related tasks can amplify perceived workload, highlighting the importance of clear communication and support to minimise burden and enhance overall care experience.

### Comparison with Previous Work

We found that patients’ experiences with real-time access to ECG device monitoring data varied, with reports of emotions such as reassurance and anxiety, stemming from perceived information needs on how to interpret the data. Consistent with earlier qualitative research on how individuals interpret personal health data, uncertainties or ambiguities in the data often trigger feelings of anxiety or confusion. ^11,27,28^ A qualitative study of cardiac patients using Fitbit data showed that while patients gained valuable insights, they also felt uncertain when their physical symptoms did not align with the data.^28^ Alternatively, a qualitative study of patients’ experiences with app-based feedback from implanted cardiac monitors found that patients found it easier to cope by verifying whether their symptoms aligned with their data, often assuming that "no news was good news" and expecting clinicians to reach out when necessary.^11^ Qualitative studies on consumer health devices suggest that offering regular reassurance, through additional actionable feedback within linked smartphone apps, can increase awareness of possible health deterioration^29^ and motivate health behaviours^30^. Given these varied emotions, providing education and involving patients in shared decision-making about data access preferences are essential to minimise potential anxiety and empower patients to take action regarding their health, as needed.^31^

The monitoring period is a key opportunity to support patients’ digital health literacy. The limits of patients’ digital health literacy are suggested in a qualitative study on patient and clinician views on the implementation of digital technology for heart disease diagnosis, highlighting patients’ desire for simplicity and adequate initial setup support.^32^ Moreover, a systematic review and content analysis of barriers and facilitators to patient engagement with remote health-tracking technologies highlighted the importance of practical support, such as clear information, to address technical issues, a common usability challenge.^12^ These findings reflect the limited consumer-focused resources to build digital health literacy^33^, making engagement challenging. Insufficient patient education could worsen disparities by disadvantaging those with lower digital health literacy.^34^ A position paper on using digital devices to detect and manage arrhythmias recommends that clinicians consider patients’ digital health literacy when delivering necessary information about the purpose of prescribed wearables and the expected timeframes for reviewing collected health data to reduce patient anxiety.^1^ Future care models could address two aspects of digital health literacy: technical skills related to device setup and troubleshooting, and health literacy skills for interpreting ambiguous data.^35^

Our findings suggest patients may experience treatment burden when integrating diagnostic devices into their daily routines, despite accepting their diagnostic utility. Diagnostic tests are often poorly accepted when they cause discomfort and interfere with daily activities, which are aspects of treatment burden.^36^ This issue is not unique to ECG monitoring; other long-term out-of-office tests, such as Ambulatory blood pressure monitoring (ABPM), face similar challenges. A qualitative study on patients’ experiences with ABPM reported that participants found it embarrassing and uncomfortable, especially during activities outside their home, with some reporting they would not do it again.^37^ Likewise, our participants reported social discomfort when wearing the device, and issues with symptom-logging impacted their engagement. This aligns with a qualitative study showing that excessive data entry and symptom-tracking can cause frustration and disengagement with digital technologies for heart disease diagnosis.^32^ New concepts involve classifying routine data collection as patient data work to recognise the additional demands of sensor-based health data collection and app-based self-reporting, as well as the evolving roles of patients and clinicians.^38^ Further understanding of digital treatment burden could better inform device design to improve user experience and facilitate engagement.

### Strengths and Limitations

We conducted a qualitative study to explore the lived experiences of using a prescribed patch ECG monitor as part of routine care. We engaged with participants across primary and secondary care settings. The demographic characteristics of our sample adequately represent those of patients typically prescribed these devices^4,39^. Our study findings should be interpreted in the context of some limitations. Due to the recruitment approach, we were unable to specifically identify and target individuals who had premature device discontinuation or who may have opted not to provide feedback due to bad experiences. We did not capture digital health literacy levels, which may have influenced the participant’s experience with their device. Finally, we did not involve patients in piloting the interview guide, so we may have overlooked relevant patient experiences. However, the semi-structured nature of the interview allowed participants to share any additional insights or important topics.

### Implications

Our findings suggest that educational resources based on patient experiences and consideration of patients’ preferences on the provision of their data, could improve patient adherence and engagement with the device. Clinicians prescribing these devices should consider patients’ digital and health literacy levels and explain the purpose of the monitoring device and the expected timelines for data review to reduce potential anxiety during and after monitoring.^40^ Concerns around routine data work include the added burden of monitoring, as patients may assume additional roles beyond data collection.^41^ Despite the advantages of automated data collection, the additional roles patients assume in managing these data can be burdensome, particularly in the absence of accessible information and technical support^13^. Future wearable apps might feature personalised dashboards displaying actionable information, such as tailored notifications, troubleshooting support, and heart rhythm and data abnormality alerts^42^. Digital educational strategies^43^ could also support a limited workforce’s capacity to teach patients the technical and health literacy skills needed to independently manage their device and remain engaged during monitoring.^31^

## Conclusion

This qualitative study found that participants valued the prescribed patch ECG monitor as a diagnostic tool despite its perceived burden on daily activities. Participants had mixed experiences with access to their live-ECG data, with some uncertain about the process of receiving or seeking clinical feedback during or after the monitoring. Further research should explore patient perspectives on their role in engaging with their health data to inform the personalisation of wearable devices to their needs, minimise treatment burden and maximise engagement.

## Data Availability

The data that support the findings of this study are available from the corresponding author upon reasonable request.

## Acknowledgements

We acknowledge Rebecca Halliwell’s contribution to her role in facilitating recruitment efforts.

## Abbreviations

CVD: Cardiovascular Disease
GP: General Practice; General Practitioner

